# Cervicovaginal and anal self-sampling for HPV testing in a transgender and gender diverse population assigned female at birth: comfort, difficulty, and willingness to use

**DOI:** 10.1101/2023.08.15.23294132

**Authors:** Erin F Welsh, Emily C. Andrus, Claire B. Sandler, Molly B. Moravek, Daphna Stroumsa, Shanna K. Kattari, Heather M. Walline, Christine M. Goudsmit, Andrew F. Brouwer

## Abstract

**Background:** Transgender and gender diverse (TGD) people assigned female at birth (AFAB) face numerous barriers to preventive care, including for HPV and cervical cancer screening. Self-sampling options may expand access to HPV testing for TGD people AFAB.

**Methods:** We recruited TGD individuals AFAB to collect cervicovaginal and anal specimens at-home using self-sampling for HPV testing, and individuals reported their perceptions of self-sampling. Associations between demographic and health characteristics and each of comfort of use, ease of use, and willingness to use self-sampling were estimated using robust Poisson regression.

**Results:** The majority of the 101 participants who completed the study reported that the cervicovaginal self-swab was not uncomfortable (68.3%) and not difficult to use (86.1%), and nearly all (96.0%) were willing to use the swab in the future. Fewer participants found the anal swab to not be uncomfortable (47.5%), but most participants still found the anal swab to not be difficult to use (70.2%) and were willing to use the swab in the future (89.1%). Participants were more willing to use either swab if they had not seen a medical professional in the past year. About 70% of participants who reported negative experiences with either self-swab were still willing to use that swab in the future.

**Conclusions:** TGD AFAB individuals were willing to use and preferred self-sampling methods for cervicovaginal and anal HPV testing. Developing clinically approved self-sampling options for cancer screening could expand access to HPV screening for TGD AFAB populations.

## Introduction

The human papillomavirus (HPV) is the most common STI among sexually active people, with most adults contracting at least one HPV strain during their lifetime.^1^ HPV infections are typically symptomatic and clear in 1–2 years, but they have the potential to contribute to future health issues such as genital warts and cancer of the cervix, anal canal, and oropharynx. Approximately 22,000 instances of HPV-attributable cancer are diagnosed annually in the US among those assigned female at birth (AFAB), including 11,100 cases of cervical cancer, 4,700 cases of anal cancer, and 2,300 cases of oropharyngeal cancer each year.^1^ While the risk of HPV transmission can be reduced by using protection during sexual activity, these measures do not mitigate risk completely. Additionally, while HPV vaccines are highly effective in unexposed individuals, uptake has lagged, and the vaccines are less effective in individuals who have been previously infected.^2^ Accordingly, cervical cancer screening remains an important preventive measure.

Per the US Preventive Services Task Force (USPSTF), current cervical cancer screening guidelines recommend that (cisgender) women aged 21–29 years should undergo a Pap smear every three years. For women aged 30–65 years, the USPSTF suggests either continuing with a Pap smear every three years, or combining it with high-risk HPV testing, which should be conducted every five years.^3^ Preventive cancer screening relies on regular engagement with health care practitioners and facilities. Transgender and gender diverse (TGD) individuals AFAB face many barriers to receiving preventive care, including cancer screening. (In this study, TGD includes transgender, nonbinary/gender diverse, both, or, in the case of AFAB individuals identifying as male, neither identities). TGD individuals are an especially vulnerable population, facing numerous barriers to accessing healthcare services, including discrimination, lack of provider knowledge and training, insurance exclusions, and financial barriers.^4,5^ Many TGD individuals report delaying medical care due to fear of discrimination, providers lacking trans specific medical care knowledge, dysphoria, and discomfort.^4,5^ Delaying medical care places additional health risks on an already vulnerable population. Hysterectomy is rare among TGD AFAB individuals^6^, so cervical screening remains an important part of preventive care for this population. HPV testing and cervical cancer screening recommendations do not distinguish between cisgender (i.e., non-TGD) women and TGD individuals, despite evidence that TGD patients are substantially more likely to have inconclusive test results.^7,8^ In addition, healthcare providers may recommend anal cancer screening using anal Pap smears or HPV testing in transmasculine individuals who engage in receptive anal sex.^9^

There is limited research on the prevalence of HPV and HPV-related cancers in transmasculine individuals compared to the cisgender population.^10^ While TGD individuals AFAB are often considered by providers and patients to be at lower risk of HPV infection because of supposedly lower rates of intercourse with partners with a penis, some studies suggest that transmasculine individuals may have a higher risk of cervical cancer and other HPV-related cancers than administering self-collected cervicovaginal and anal swabs for HPV testing at home, as well as swab comfort, ease-of-use, and acceptability.

## Materials and Methods

### Study design and population

This observational study enrolled participants in 2020–21 and investigated prevalence of cervicovaginal, oral, and anal HPV among TGD people AFAB using at-home self-sampling and to assess comfort, ease-of-use, and preference for at-home cervicovaginal and anal HPV testing.^15^ This analysis focuses on participant’s perceptions and experience of the at-home cervicovaginal and anal specimen sampling process. The University of Michigan Institutional Review Board and the Rogel Cancer Center Protocol Review Committee approved consent documents and study protocol (HUM00166980). Ten TGD community members reviewed the study protocol, surveys, and sample collection instructions; each person was compensated with a $100 gift card for their time.

Participants were recruited through the Michigan Medicine Comprehensive Gender Services Program, the Michigan Medicine research study recruitment website, social media posts, outreach to LGBT+ organizations, and word of mouth referrals. To qualify for the study, participants had to meet the following inclusion criteria: (1) be able and willing to provide consent, (2) be between the ages of 21–65 years of age, (3) be assigned female at birth, (4) identify as male, masculine, nonbinary, or another TGD identity, and (5) live in Michigan, Illinois, Indiana, Ohio, or Wisconsin. Individuals were excluded from participating if they: (1) had their cervix removed, or (2) self-reported being pregnant. Samples could not be collected if participants were menstruating. Participants gave written, informed consent virtually, using the SignNow platform. Participants were compensated with a $50 gift card for full study participation.

### Procedures

#### Sample self-collection

Individuals who consented to participate were mailed a study test kit that included materials and instructions for the collection of cervicovaginal, oral, and anal biospecimens. Participants were asked to provide an oral rinse sample using Scope brand mouthwash (Proctor & Gamble), a cervicovaginal sample using an Evelyn Brush (Rovers Medical Devices), and an anal sample using a Dacron flocked swab (Puritan Medical Products).^16,17^ The Evalyn Brush is a cervicovaginal sampling device designed to collect cells near the cervix. The protocol for anal swab sampling was adapted from previously published protocols.^18,19^ More details on sample collection and analysis are provided in McIntosh et al.^15^

#### Questionnaire

Following sample collection, participants were asked to complete two questionnaires. First, participants completed a questionnaire adapted from the validated instrument developed for the Michigan HPV and Oropharyngeal Cancer Study, consisting of questions relating to demographic information, including age, race/ethnicity, and socioeconomic status; behavior, including alcohol, tobacco, and other drug use; and sexual practices, sexual history, and sexual health care (including HPV vaccination status), testosterone status, general health, and history of STIs.^19^ Secondly, to assess participants’ individual at-home self-sampling experiences, participants answered a questionnaire was adapted from a previously developed instrument, consisting of the comfort, ease-of-use, and preference for the cerivcovaginal and anal self-collection procedures.^20^ Oral sampling for HPV was not similarly assessed as it uses a non-invasive sampling method (oral rinse).

### Statistical analysis

This study assessed three primary outcomes: perception of self-sampling swab comfortability, perception of self-sampling difficulty, and willingness to use the self-sampling method for future HPV testing. Comfort and ease-of-use were asked on a Likert scale (1-5) but analyzed as binary outcomes with values 4-5 indicating the swab was uncomfortable or difficult to use. Outcomes were assessed for both the cervicovaginal and anal self-sampling experiences. Prevalence ratios and 95% confidence intervals were estimated using Poisson regression with robust error variance. All analyses were conducted using SAS 9.4. Statistical significance is reported at level 1Z = 0.05.

## Results

### Sample characteristics

We enrolled 137 TGD participants, and 101 participants completed both the HPV self-swab collections and questionnaires. The mean age of the cohort was 28.6 years, with two-thirds of the population falling into the 21–29-year age group (Table 1). There was an even distribution of gender identity between those identifying as nonbinary, genderfluid, agender, or another gender diverse identity (48.0%) and as transgender male, transmasculine, male, or another transgender identity (49.0%). Most of the participants identified their sexual orientation as either bisexual, pansexual, or omnisexual (43.1%) or queer (36.3%). Most participants reported ever delaying medical care due to concerns relating to their gender identity (82.2%), such as expected lack of provider knowledge on transgender care, fear of discrimination, and dysphoria (Table 2). Three-quarters of participants reported having transitioned or be in the process of transitioning from their gender assigned at birth, and, of those individuals, 89.3% were currently on testosterone hormone therapy for masculinization. A small percentage reported a previous history of pregnancy (13.9%). The average BMI was 29.3, with there being an increasing distribution among individuals who are classified as normal weight (26.6%), overweight (33.0%), and obese (36.2%). Most of the population had prior experience receiving a cervical Pap test (78.2%), but few (5.9%) reported prior experience of an anal HPV test. Over half of the population reported receiving at least one dose of the HPV vaccine.

**Table 1.** Demographic characteristics of participants. (N=101)

|  | n | % |
| --- | --- | --- |
| <u>Age Group</u> |  |  |
| 21-29 | 67 | 66.7 |
| 30-39 | 29 | 28.4 |
| 40-59 | 6 | 5.9 |
| <u>Race/Ethnicity</u> |  |  |
| White | 84 | 82.3 |
| Multiracial | 8 | 7.8 |
| Asian | 5 | 4.9 |
| Black, African American, or African | 5 | 4.9 |
| <u>Gender Identity</u> |  |  |
| Nonbinary, genderfluid, agender | 49 | 48.5 |
| Transgender male / transmasculine / male | 49 | 48.5 |
| Other / Prefer not to label | 2 | 3.0 |
| <u>Sexual Orientation</u> |  |  |
| Straight or heterosexual | 3 | 2.9 |
| Gay / Lesbian | 9 | 8.8 |
| Bisexual, pansexual, or omnisexual | 44 | 43.1 |
| Asexual or demisexual | 8 | 7.8 |
| Queer | 37 | 36.3 |
| Other / Prefer not to label | 1 | 1.1 |
| <u>Highest Level of Education</u> |  |  |
| High school or less | 8 | 7.8 |
| Some college | 27 | 26.5 |
| 2- or 4-year degree | 46 | 45.1 |
| Professional or doctorate degree | 21 | 20.6 |
| <u>Marital Status</u> |  |  |
| Never married or partnered | 25 | 11.8 |
| Single partner, never married | 12 | 24.5 |
| Single partner, married | 47 | 46.1 |
| Multiple committed partners | 11 | 10.9 |
| Separated, divorced, or widowed | 7 | 6.7 |
| <u>Employment status</u> |  |  |
| Student | 22 | 21.6 |
| Disabled and not working | 10 | 9.8 |
| Unemployed, not looking for work | 5 | 4.9 |
| Unemployed, looking for work | 12 | 11.8 |
| Part-time | 15 | 14.7 |
| Full-time | 38 | 37.1 |
| <u>Yearly household income</u> |  |  |
| Less than \$10,000 | 17 | 16.8 |
| \$10,000 - \$19,999 | 19 | 18.8 |
| \$20,000 - \$39,999 | 20 | 19.9 |
| \$40,000 - \$49,999 | 21 | 20.8 |
| \$50,000 + | 24 | 23.8 |
| <u>Developed Environment</u> |  |  |
| Rural | 6 | 5.9 |
| Suburban | 49 | 48.0 |
| Urban | 47 | 46.1 |
| <u>Medical Insurance</u> |  |  |
| Public insurance | 31 | 30.7 |
| Private insurance | 67 | 66.3 |
| None/Unsure | 3 | 3.0 |

**Table 2.** Medical and sexual history of participants. (N=101)

|  | n | % |
| --- | --- | --- |
| <u>Body Mass Index (BMI), kg/m<sup>2</sup> (N=94)</u> |  |  |
| Underweight (< 18.5 kg/m<sup>2</sup>) | 4 | 4.3 |
| Normal weight (18.5-24.9 kg/m<sup>2</sup>) | 25 | 26.6 |
| Overweight (25.0-29.9 kg/m<sup>2</sup>) | 31 | 33.0 |
| Obese (≥ 30 kg/m<sup>2</sup>) | 34 | 36.2 |
| <u>Visited a medical professional within the past year</u> |  |  |
| No | 8 | 7.9 |
| Yes | 93 | 82.1 |
| <u>Number of lifetime sexual partners</u> |  |  |
| < 5 | 34 | 33.7 |
| 5-10 | 23 | 22.8 |
| > 10 | 44 | 43.6 |
| <u>Number of sexual partners within the past 6 months (N=99)</u> |  |  |
| 0 | 19 | 19.2 |
| 1 | 60 | 60.6 |
| 2-4 | 17 | 17.2 |
| 5+ | 3 | 3.0 |
| <u>History of vaginal/ front hole sex (N=97)</u> |  |  |
| No | 2 | 2.1 |
| Yes | 95 | 97.9 |
| <u>History of anal sex (N=95)</u> |  |  |
| No | 38 | 40.0 |
| Yes | 57 | 60.0 |
| <u>History of any STI diagnosis</u> |  |  |
| No | 77 | 76.2 |
| Yes | 24 | 23.8 |
| <u>Transitioned/transitioning from gender assigned at birth</u> |  |  |
| No | 25 | 24.8 |
| Yes | 76 | 75.2 |
| <u>Current hormone usage for masculinization (N=57)</u> |  |  |
| No | 6 | 10.5 |
| Yes | 51 | 89.5 |
| <u>History of pregnancy</u> |  |  |
| No | 87 | 86.1 |
| Yes | 14 | 13.9 |
| <u>Ever had cervical Pap test or HPV test</u> |  |  |
| No | 22 | 21.8 |
| Yes | 79 | 78.2 |
| <u>Have had prior anal Pap test</u> |  |  |
| No | 95 | 94.1 |
| Yes | 6 | 5.9 |
| <u>Ever delayed checkups</u> |  |  |
| No | 18 | 17.8 |
| Yes, provider lacked knowledge on trans-specific care | 6 | 6.0 |
| Yes, fear of discrimination | 16 | 15.8 |
| Yes, to avoid dysphoria or physical discomfort | 61 | 60.4 |
| <u>Have received any shots of the HPV vaccine</u> |  |  |
| No | 43 | 42.6 |
| Yes | 58 | 57.4 |

### Perceptions of cervicovaginal self-swab collection

Overall, perceptions of the cervicovaginal self-swab collection for HPV were positive (Table 3). A large proportion of the subjects reported that using the cervicovaginal swab was not uncomfortable (68.3%), as well as not difficult to use (86.1%). Participants with a history of pregnancy were more likely to report that the swab was not uncomfortable (PR 1.31; 95% CI 1.91, 1.70), and those who reported delayed medical care due to their gender identity were less likely to report the swab was not uncomfortable compared to those who never delayed medical care (PR 0.72; 95% CI 0.56, 0.92). None of the demographic or health-related variables were significantly associated with an individual’s perception that the cervicovaginal swab was easy to use. Perceptions of the swab being comfortable and being easy to use were strongly correlated.

**Table 3.**
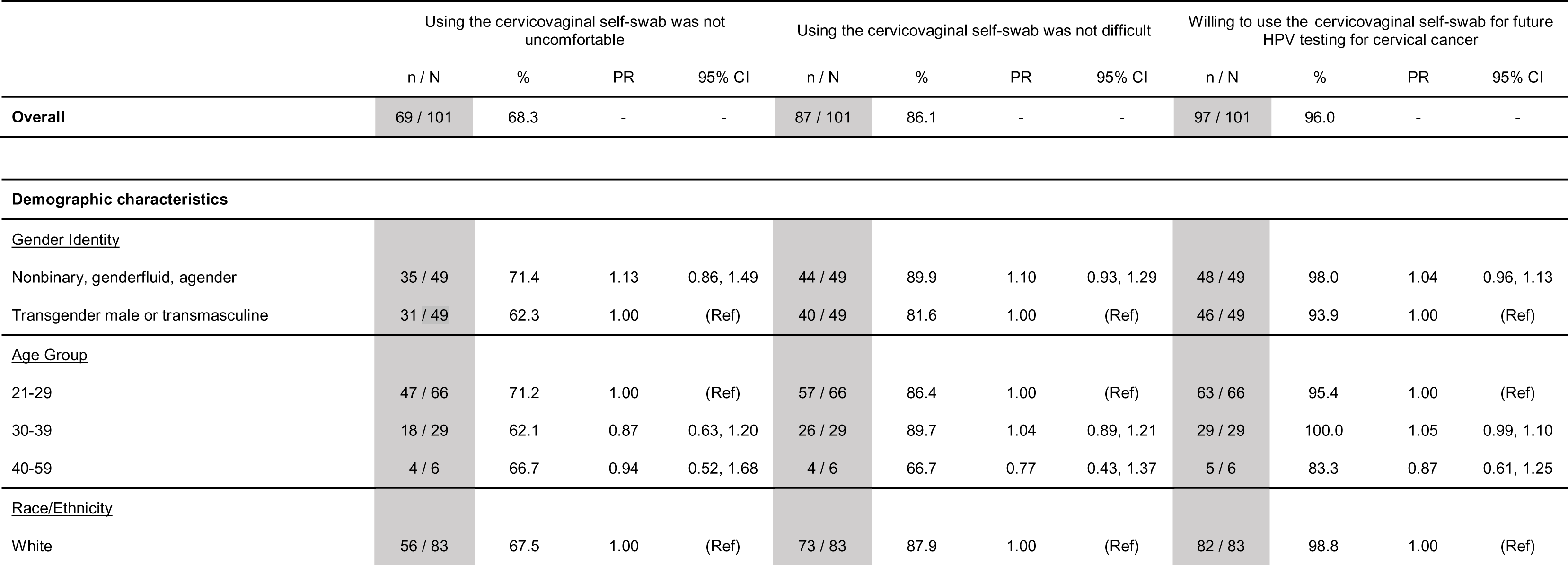
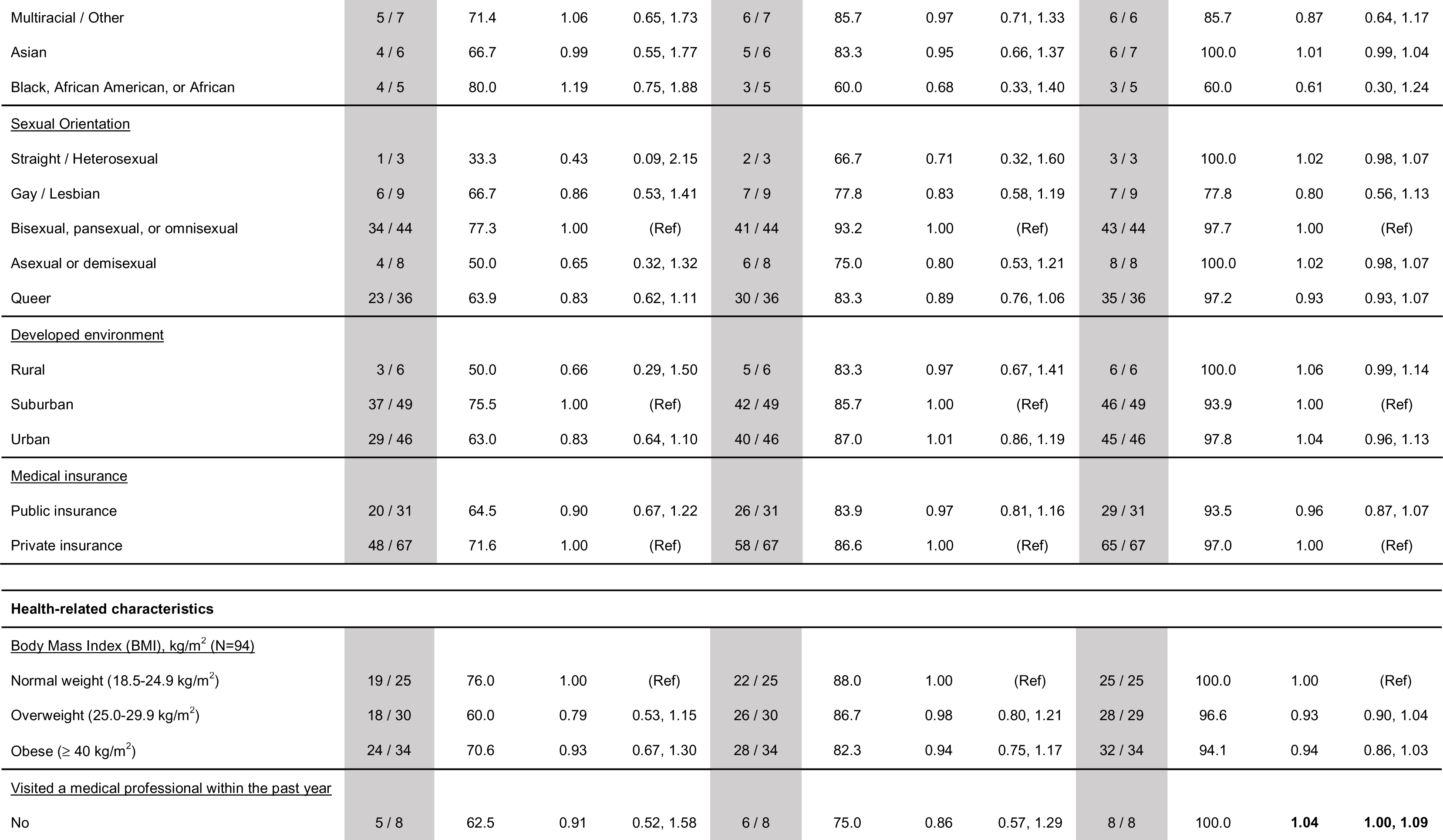
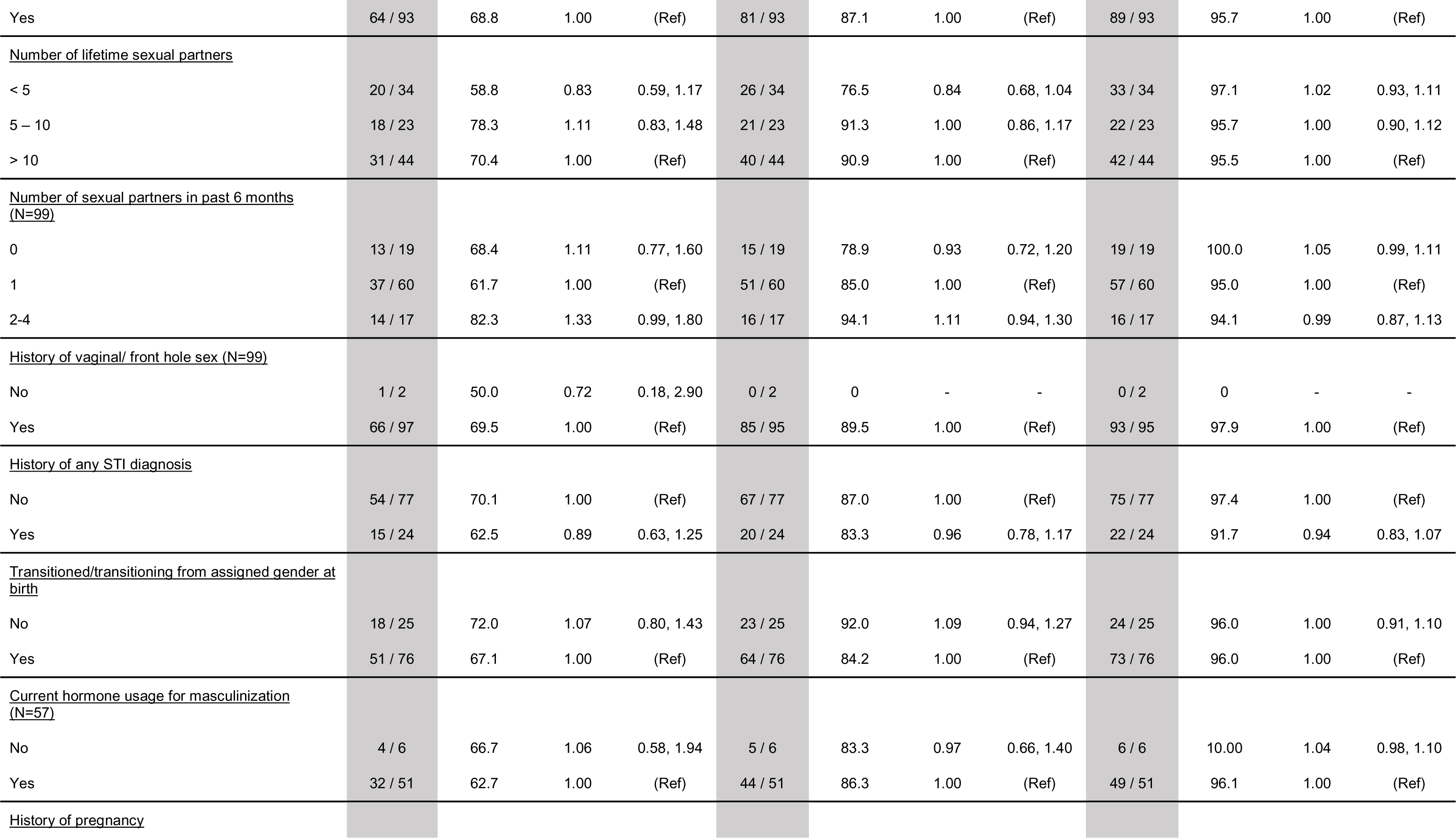
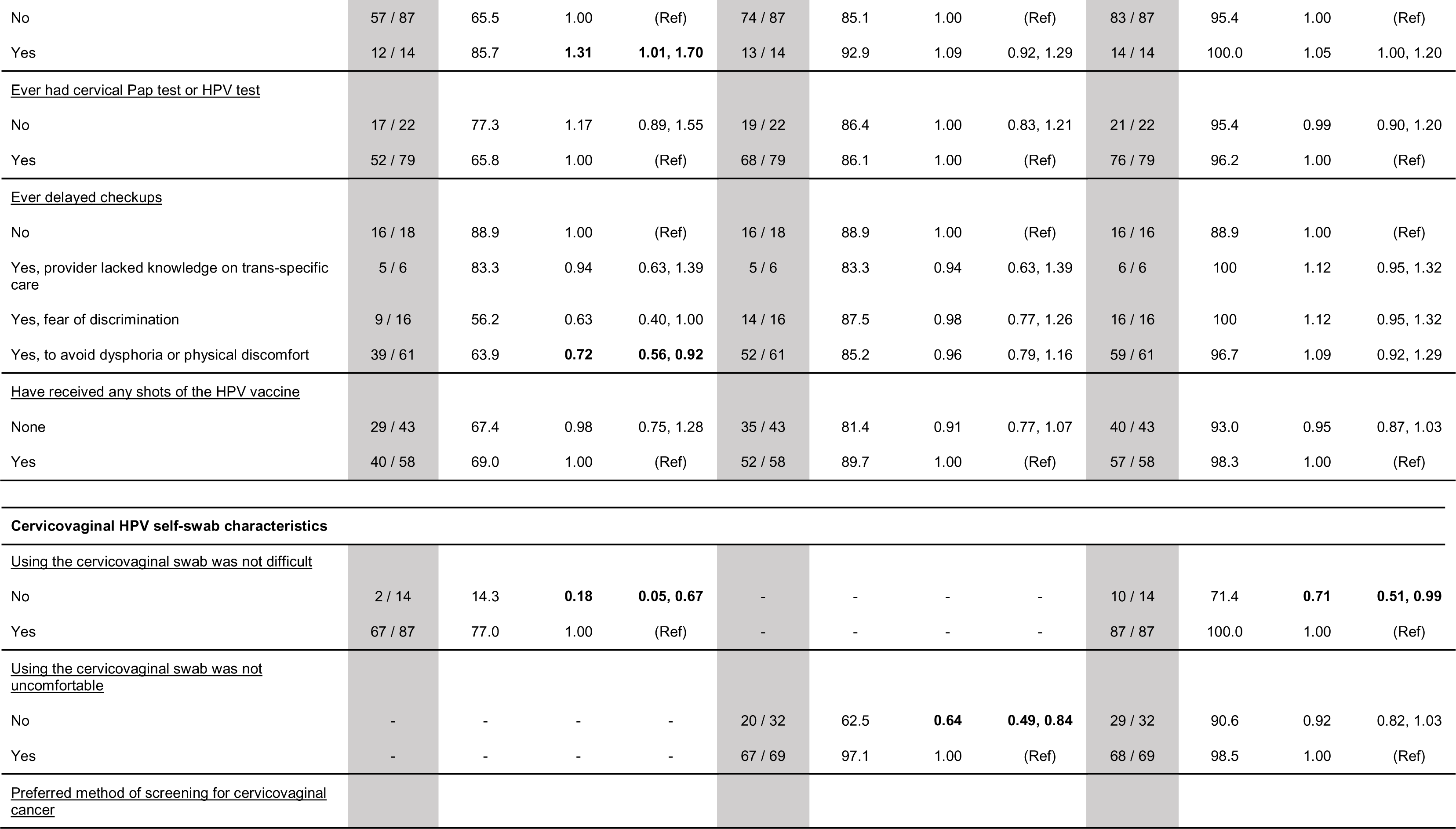
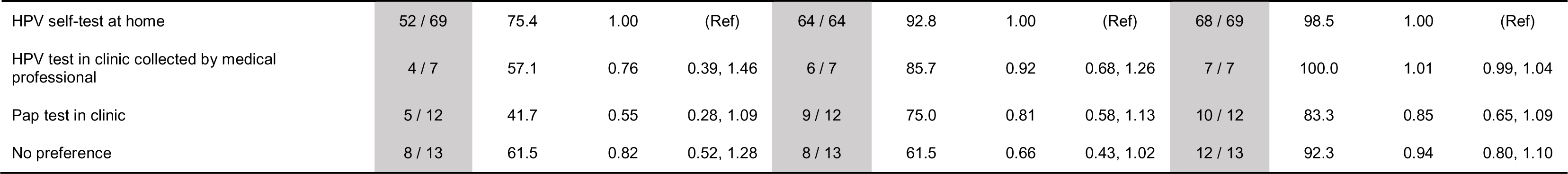
Prevalence and prevalence ratios of participant perceptions that using the cervicovaginal self-swab was not uncomfortable, using the cervicovaginal self-swab was not difficult, and being willing to use the cervicovaginal self-swab again for future HPV testing for cervical cancer by participant demographics, medical characteristics, and cross comparisons to the cervicovaginal self-swab characteristics and outcomes. Prevalence ratios found to be statistically significant are bolded. n = number of individuals who responded ‘yes’ within group, N = denominator for the given category, % = prevalence of individuals who responded ‘yes’ within each group (reported as a percentage), PR = prevalence ratio of individuals who responded ‘yes’ in each group compared to the referent group, and 95% CI = 95% confidence interval for each prevalence ratio. Statistics were calculated using Poisson regression with robust error variance.

Nearly all participants expressed that they would be willing to use the cervicovaginal self-swab in the future for HPV and cervical cancer screening (96.0%). There were few significant differences in willingness to use the self-swab by any of the demographic, sexual history, medical history, or transition history characteristics, except that those who had not seen a medical professional within the past year were more willing to use the cervicovaginal self-swab than those that had (PR 1.04; 95% CI 1.00, 1.09). Willingness to use the swab was lower among those who found the swab to be uncomfortable (PR 0.71; 95% CI 0.51, 0.99). There was an overall preference for at-home self-swab collection for cervicovaginal HPV testing, with 68.3% preferring at-home testing overall, compared to 6.9% preferring in clinic collection by a physician, 11.9% preferring a pap test in the clinic, and 12.9% with no preference.

### Perceptions of anal self-swab collection

Perceptions of the anal self-swab for HPV testing were less positive than those about the cervicovaginal self-swab (Table 4). Only half (47.5%) of participants reported the anal swab to not be uncomfortable, and though most still perceived the anal swab to not be difficult to use (70.2%). Participants were more likely to find the swab to be uncomfortable if they had public health insurance compared to private insurance (PR 0.51; 95% CI 0.28, 0.92), but were more likely to find the swab to not be uncomfortable if they had not seen a medical professional within the past year compared to those who had (PR 1.66; 95% CI 1.05, 2.63), had delayed medical care due to the provider lacking trans-specific knowledge compared to those that had not (PR 3.00; 95% CI 1.07, 8.43), or delayed medical care due to fear of discrimination compared to those that had not (PR 2.81; 95% CI 1.09, 7.23). Participants were more likely to find the swab to be difficult to use if they were 30–39 years old compared to 21–29 years old (PR 0.66; 95% CI 0.45, 0.95), but more likely to not find the swab difficult to use if they were Asian compared to White (PR 1.43; 95% CI 1.24, 1.65), lived in a rural area compared to a suburban area (PR 1.53; 95% CI 1.25, 1.88), delayed medical care because the medical professional lacked knowledge on trans-specific healthcare compared to those that did not (PR 2.00; 95% CI 1.26, 1.67). Comfort and ease-of-use of the swab were again strongly correlated.

**Table 4.**
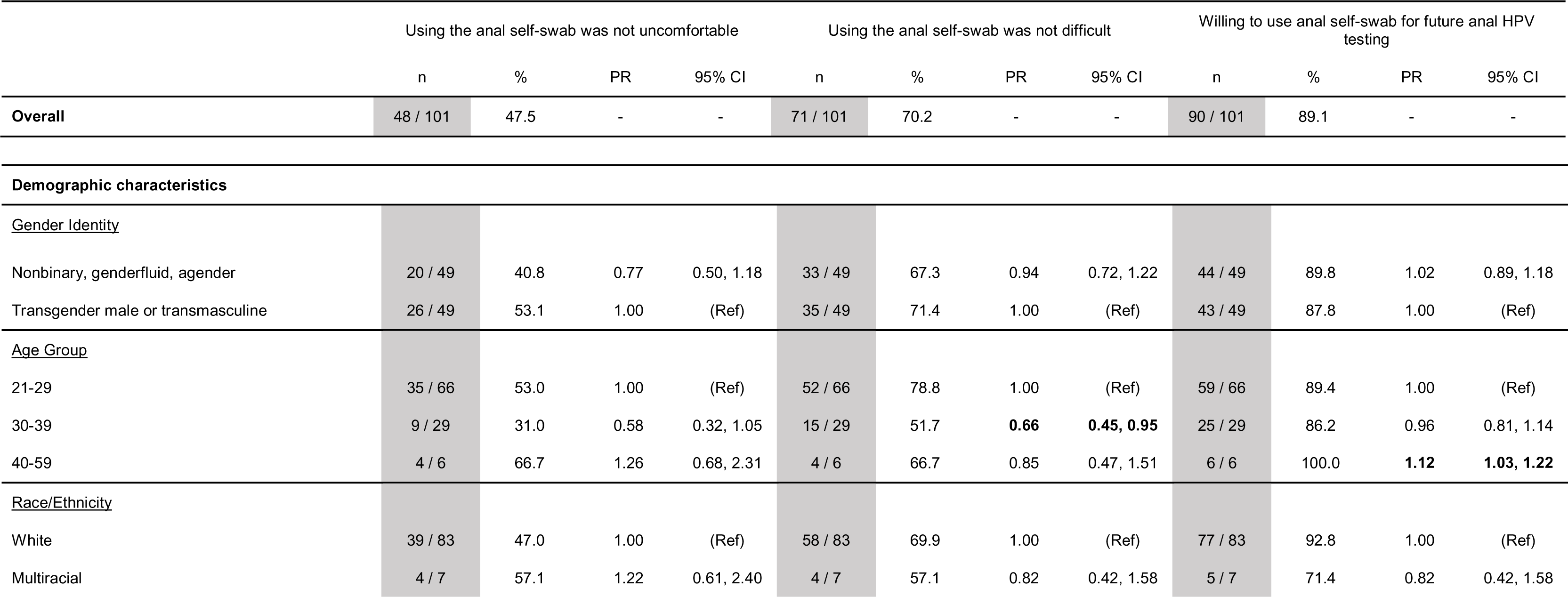
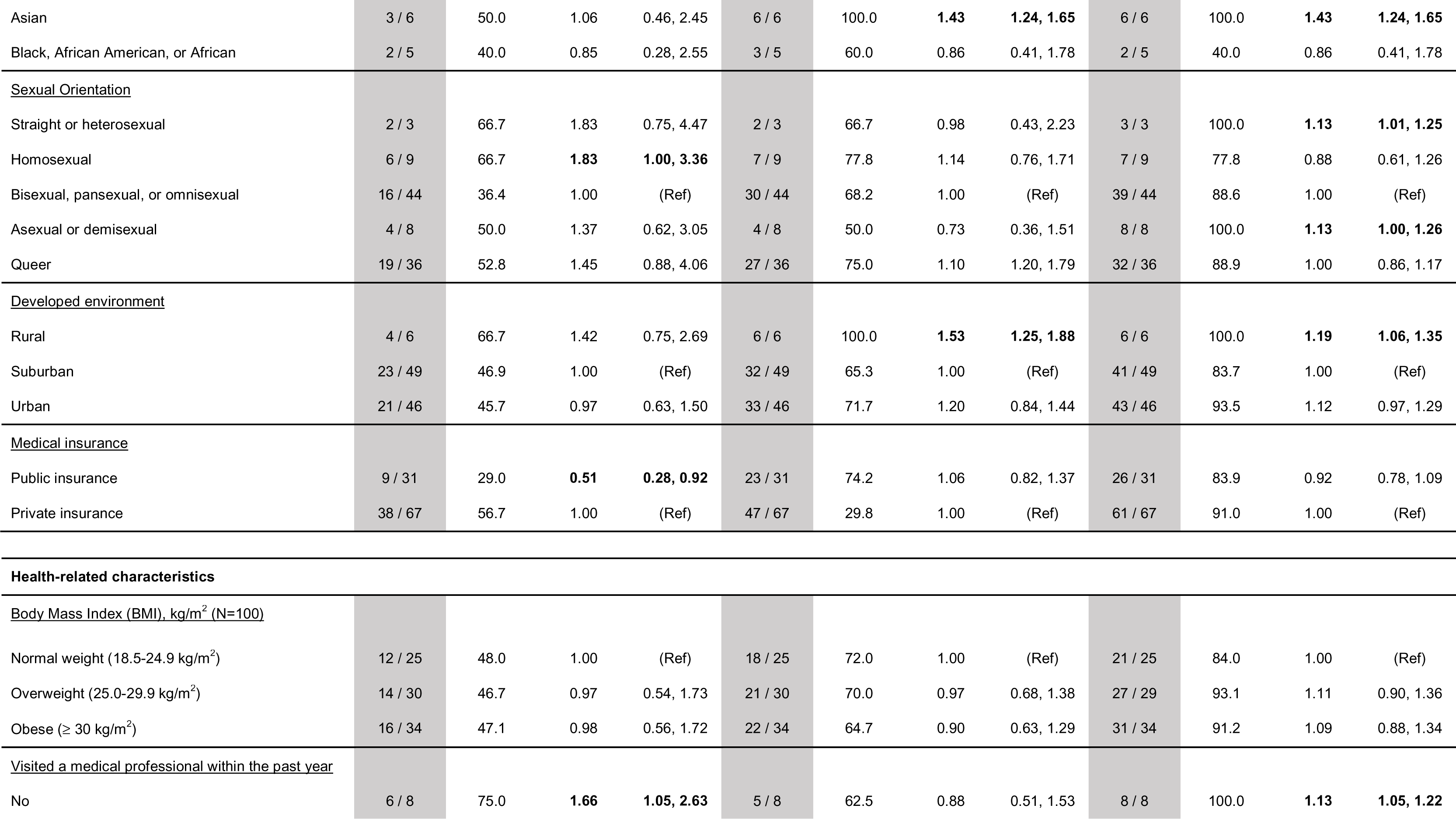
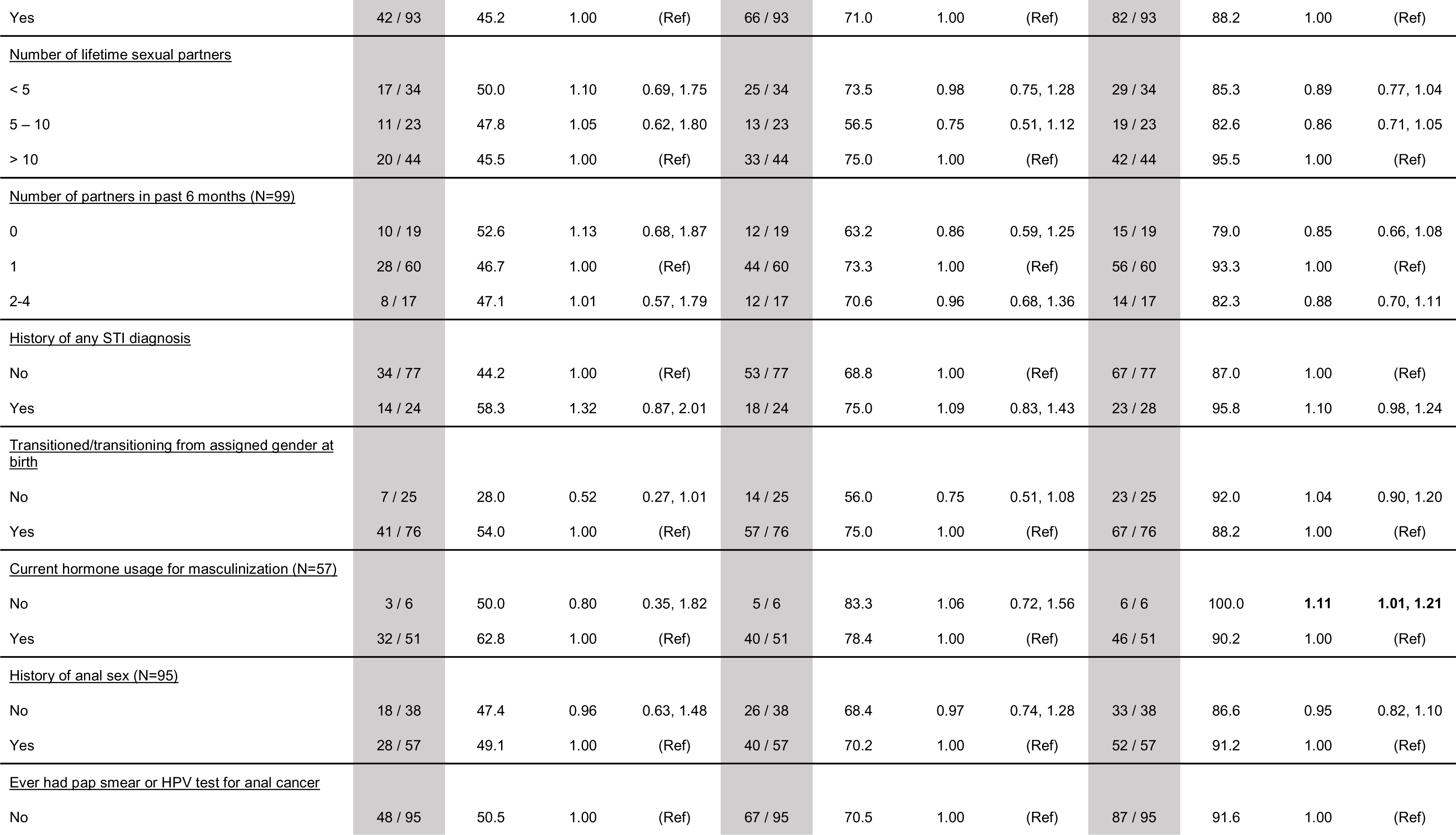
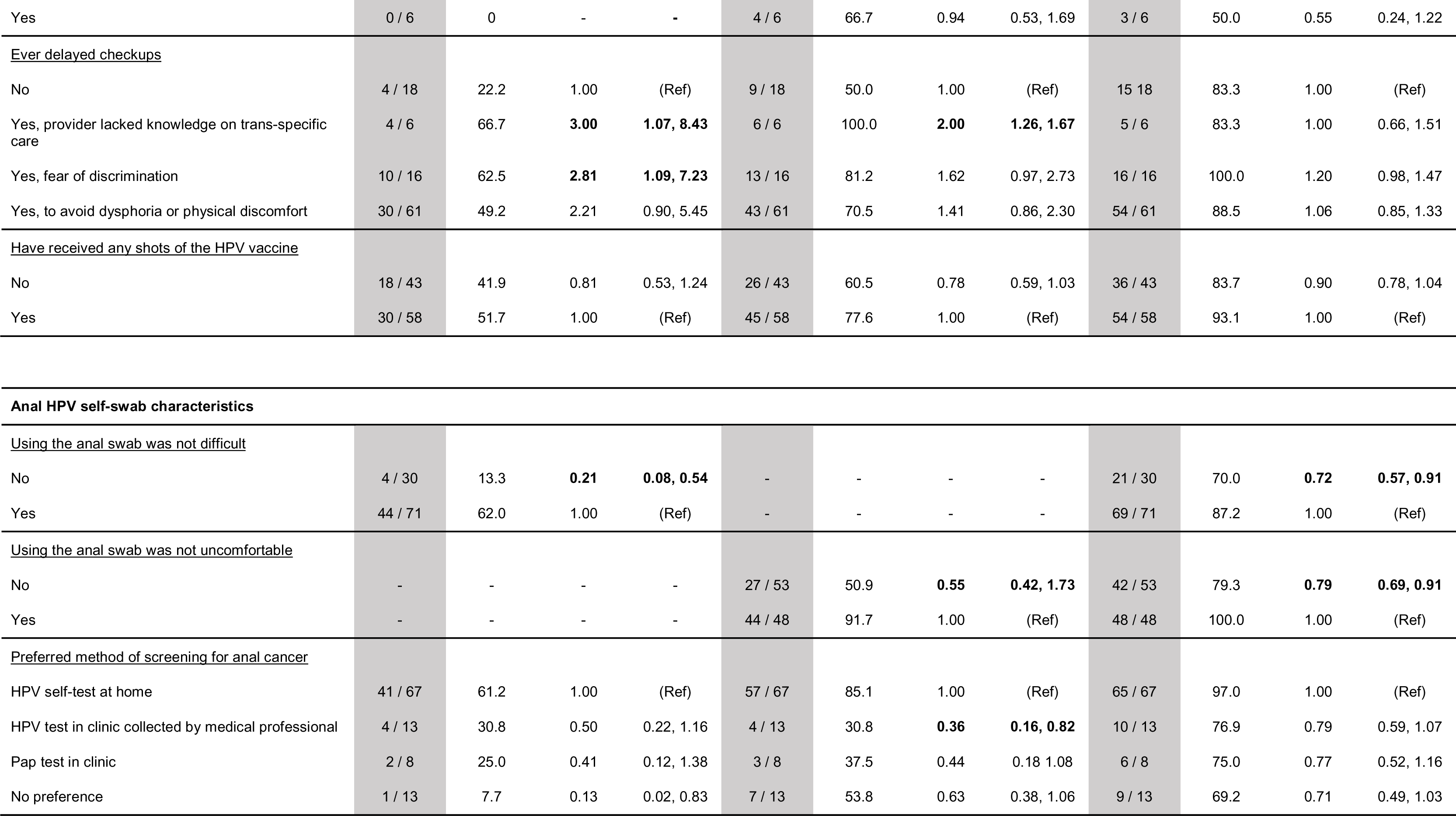
Prevalence and prevalence ratios of participant perceptions that using the anal self-swab was not uncomfortable, using the anal self-swab was not difficult, and being willing to use the anal self-swab again for future HPV testing for anal cancer by participant demographics, medical characteristics, and cross comparisons to the anal self-swab characteristics and outcomes. Prevalence ratios found to be statistically significant are bolded. n = number of individuals who responded ‘yes’ within group, N = denominator within the given category, % = prevalence of individuals who responded ‘yes’ within each group (reported as a percentage), PR = prevalence ratio of individuals who responded ‘yes’ in each group compared to the referent group, and 95% CI = 95% confidence interval for each prevalence ratio. Statistics were calculated using Poisson regression with robust error variance.

|  | Using the anal self-swab was not uncomfortable |  |  |  | Using the anal self-swab was not difficult |  |  |  | Willing to use anal self-swab for future anal HPV testing |  |  |  |
| --- | --- | --- | --- | --- | --- | --- | --- | --- | --- | --- | --- | --- |
|  | n | % | PR | 95% CI | n | % | PR | 95% CI | n | % | PR | 95% CI |
| Overall | 48 / 101 | 47.5 | - | - | 71 / 101 | 70.2 | - | - | 90 / 101 | 89.1 | - | - |
| Demographic characteristics |  |  |  |  |  |  |  |  |  |  |  |  |
| <u>Gender Identity</u> |  |  |  |  |  |  |  |  |  |  |  |  |
| Nonbinary, genderfluid, agender | 20 / 49 | 40.8 | 0.77 | 0.50, 1.18 | 33 / 49 | 67.3 | 0.94 | 0.72, 1.22 | 44 / 49 | 89.8 | 1.02 | 0.89, 1.18 |
| Transgender male or transmasculine | 26 / 49 | 53.1 | 1.00 | (Ref) | 35 / 49 | 71.4 | 1.00 | (Ref) | 43 / 49 | 87.8 | 1.00 | (Ref) |
| <u>Age Group</u> |  |  |  |  |  |  |  |  |  |  |  |  |
| 21-29 | 35 / 66 | 53.0 | 1.00 | (Ref) | 52 / 66 | 78.8 | 1.00 | (Ref) | 59 / 66 | 89.4 | 1.00 | (Ref) |
| 30-39 | 9 / 29 | 31.0 | 0.58 | 0.32, 1.05 | 15 / 29 | 51.7 | **0.66** | **0.45, 0.95** | 25 / 29 | 86.2 | 0.96 | 0.81, 1.14 |
| 40-59 | 4 / 6 | 66.7 | 1.26 | 0.68, 2.31 | 4 / 6 | 66.7 | 0.85 | 0.47, 1.51 | 6 / 6 | 100.0 | **1.12** | **1.03, 1.22** |
| <u>Race/Ethnicity</u> |  |  |  |  |  |  |  |  |  |  |  |  |
| White | 39 / 83 | 47.0 | 1.00 | (Ref) | 58 / 83 | 69.9 | 1.00 | (Ref) | 77 / 83 | 92.8 | 1.00 | (Ref) |
| Multiracial | 4 / 7 | 57.1 | 1.22 | 0.61, 2.40 | 4 / 7 | 57.1 | 0.82 | 0.42, 1.58 | 5 / 7 | 71.4 | 0.82 | 0.42, 1.58 |
| Asian | 3 / 6 | 50.0 | 1.06 | 0.46, 2.45 | 6 / 6 | 100.0 | 1.43 | 1.24, 1.65 | 6 / 6 | 100.0 | 1.43 | 1.24, 1.65 |
| Black, African American, or African | 2 / 5 | 40.0 | 0.85 | 0.28, 2.55 | 3 / 5 | 60.0 | 0.86 | 0.41, 1.78 | 2 / 5 | 40.0 | 0.86 | 0.41, 1.78 |
| <u>Sexual Orientation</u> |  |  |  |  |  |  |  |  |  |  |  |  |
| Straight or heterosexual | 2 / 3 | 66.7 | 1.83 | 0.75, 4.47 | 2 / 3 | 66.7 | 0.98 | 0.43, 2.23 | 3 / 3 | 100.0 | 1.13 | 1.01, 1.25 |
| Homosexual | 6 / 9 | 66.7 | 1.83 | 1.00, 3.36 | 7 / 9 | 77.8 | 1.14 | 0.76, 1.71 | 7 / 9 | 77.8 | 0.88 | 0.61, 1.26 |
| Bisexual, pansexual, or omnisexual | 16 / 44 | 36.4 | 1.00 | (Ref) | 30 / 44 | 68.2 | 1.00 | (Ref) | 39 / 44 | 88.6 | 1.00 | (Ref) |
| Asexual or demisexual | 4 / 8 | 50.0 | 1.37 | 0.62, 3.05 | 4 / 8 | 50.0 | 0.73 | 0.36, 1.51 | 8 / 8 | 100.0 | 1.13 | 1.00, 1.26 |
| Queer | 19 / 36 | 52.8 | 1.45 | 0.88, 4.06 | 27 / 36 | 75.0 | 1.10 | 1.20, 1.79 | 32 / 36 | 88.9 | 1.00 | 0.86, 1.17 |
| <u>Developed environment</u> |  |  |  |  |  |  |  |  |  |  |  |  |
| Rural | 4 / 6 | 66.7 | 1.42 | 0.75, 2.69 | 6 / 6 | 100.0 | 1.53 | 1.25, 1.88 | 6 / 6 | 100.0 | 1.19 | 1.06, 1.35 |
| Suburban | 23 / 49 | 46.9 | 1.00 | (Ref) | 32 / 49 | 65.3 | 1.00 | (Ref) | 41 / 49 | 83.7 | 1.00 | (Ref) |
| Urban | 21 / 46 | 45.7 | 0.97 | 0.63, 1.50 | 33 / 46 | 71.7 | 1.20 | 0.84, 1.44 | 43 / 46 | 93.5 | 1.12 | 0.97, 1.29 |
| <u>Medical insurance</u> |  |  |  |  |  |  |  |  |  |  |  |  |
| Public insurance | 9 / 31 | 29.0 | 0.51 | 0.28, 0.92 | 23 / 31 | 74.2 | 1.06 | 0.82, 1.37 | 26 / 31 | 83.9 | 0.92 | 0.78, 1.09 |
| Private insurance | 38 / 67 | 56.7 | 1.00 | (Ref) | 47 / 67 | 29.8 | 1.00 | (Ref) | 61 / 67 | 91.0 | 1.00 | (Ref) |
| **Health-related characteristics** |  |  |  |  |  |  |  |  |  |  |  |  |
| <u>Body Mass Index (BMI), kg/m<sup>2</sup> (N=100)</u> |  |  |  |  |  |  |  |  |  |  |  |  |
| Normal weight (18.5-24.9 kg/m<sup>2</sup>) | 12 / 25 | 48.0 | 1.00 | (Ref) | 18 / 25 | 72.0 | 1.00 | (Ref) | 21 / 25 | 84.0 | 1.00 | (Ref) |
| Overweight (25.0-29.9 kg/m<sup>2</sup>) | 14 / 30 | 46.7 | 0.97 | 0.54, 1.73 | 21 / 30 | 70.0 | 0.97 | 0.68, 1.38 | 27 / 29 | 93.1 | 1.11 | 0.90, 1.36 |
| Obese (≥ 30 kg/m<sup>2</sup>) | 16 / 34 | 47.1 | 0.98 | 0.56, 1.72 | 22 / 34 | 64.7 | 0.90 | 0.63, 1.29 | 31 / 34 | 91.2 | 1.09 | 0.88, 1.34 |
| <u>Visited a medical professional within the past year</u> |  |  |  |  |  |  |  |  |  |  |  |  |
| No | 6 / 8 | 75.0 | 1.66 | 1.05, 2.63 | 5 / 8 | 62.5 | 0.88 | 0.51, 1.53 | 8 / 8 | 100.0 | 1.13 | 1.05, 1.22 |
| Yes | 42 / 93 | 45.2 | 1.00 | (Ref) | 66 / 93 | 71.0 | 1.00 | (Ref) | 82 / 93 | 88.2 | 1.00 | (Ref) |
| <u>Number of lifetime sexual partners</u> |  |  |  |  |  |  |  |  |  |  |  |  |
| < 5 | 17 / 34 | 50.0 | 1.10 | 0.69, 1.75 | 25 / 34 | 73.5 | 0.98 | 0.75, 1.28 | 29 / 34 | 85.3 | 0.89 | 0.77, 1.04 |
| 5 – 10 | 11 / 23 | 47.8 | 1.05 | 0.62, 1.80 | 13 / 23 | 56.5 | 0.75 | 0.51, 1.12 | 19 / 23 | 82.6 | 0.86 | 0.71, 1.05 |
| > 10 | 20 / 44 | 45.5 | 1.00 | (Ref) | 33 / 44 | 75.0 | 1.00 | (Ref) | 42 / 44 | 95.5 | 1.00 | (Ref) |
| <u>Number of partners in past 6 months (N=99)</u> |  |  |  |  |  |  |  |  |  |  |  |  |
| 0 | 10 / 19 | 52.6 | 1.13 | 0.68, 1.87 | 12 / 19 | 63.2 | 0.86 | 0.59, 1.25 | 15 / 19 | 79.0 | 0.85 | 0.66, 1.08 |
| 1 | 28 / 60 | 46.7 | 1.00 | (Ref) | 44 / 60 | 73.3 | 1.00 | (Ref) | 56 / 60 | 93.3 | 1.00 | (Ref) |
| 2-4 | 8 / 17 | 47.1 | 1.01 | 0.57, 1.79 | 12 / 17 | 70.6 | 0.96 | 0.68, 1.36 | 14 / 17 | 82.3 | 0.88 | 0.70, 1.11 |
| <u>History of any STI diagnosis</u> |  |  |  |  |  |  |  |  |  |  |  |  |
| No | 34 / 77 | 44.2 | 1.00 | (Ref) | 53 / 77 | 68.8 | 1.00 | (Ref) | 67 / 77 | 87.0 | 1.00 | (Ref) |
| Yes | 14 / 24 | 58.3 | 1.32 | 0.87, 2.01 | 18 / 24 | 75.0 | 1.09 | 0.83, 1.43 | 23 / 28 | 95.8 | 1.10 | 0.98, 1.24 |
| <u>Transitioned/transitioning from assigned gender at birth</u> |  |  |  |  |  |  |  |  |  |  |  |  |
| No | 7 / 25 | 28.0 | 0.52 | 0.27, 1.01 | 14 / 25 | 56.0 | 0.75 | 0.51, 1.08 | 23 / 25 | 92.0 | 1.04 | 0.90, 1.20 |
| Yes | 41 / 76 | 54.0 | 1.00 | (Ref) | 57 / 76 | 75.0 | 1.00 | (Ref) | 67 / 76 | 88.2 | 1.00 | (Ref) |
| <u>Current hormone usage for masculinization (N=57)</u> |  |  |  |  |  |  |  |  |  |  |  |  |
| No | 3 / 6 | 50.0 | 0.80 | 0.35, 1.82 | 5 / 6 | 83.3 | 1.06 | 0.72, 1.56 | 6 / 6 | 100.0 | **1.11** | **1.01, 1.21** |
| Yes | 32 / 51 | 62.8 | 1.00 | (Ref) | 40 / 51 | 78.4 | 1.00 | (Ref) | 46 / 51 | 90.2 | 1.00 | (Ref) |
| <u>History of anal sex (N=95)</u> |  |  |  |  |  |  |  |  |  |  |  |  |
| No | 18 / 38 | 47.4 | 0.96 | 0.63, 1.48 | 26 / 38 | 68.4 | 0.97 | 0.74, 1.28 | 33 / 38 | 86.6 | 0.95 | 0.82, 1.10 |
| Yes | 28 / 57 | 49.1 | 1.00 | (Ref) | 40 / 57 | 70.2 | 1.00 | (Ref) | 52 / 57 | 91.2 | 1.00 | (Ref) |
| <u>Ever had pap smear or HPV test for anal cancer</u> |  |  |  |  |  |  |  |  |  |  |  |  |
| No | 48 / 95 | 50.5 | 1.00 | (Ref) | 67 / 95 | 70.5 | 1.00 | (Ref) | 87 / 95 | 91.6 | 1.00 | (Ref) |
| Yes | 0 / 6 | 0 | - | - | 4 / 6 | 66.7 | 0.94 | 0.53, 1.69 | 3 / 6 | 50.0 | 0.55 | 0.24, 1.22 |
| <u>Ever delayed checkups</u> |  |  |  |  |  |  |  |  |  |  |  |  |
| No | 4 / 18 | 22.2 | 1.00 | (Ref) | 9 / 18 | 50.0 | 1.00 | (Ref) | 15 / 18 | 83.3 | 1.00 | (Ref) |
| Yes, provider lacked knowledge on trans-specific care | 4 / 6 | 66.7 | **3.00** | **1.07, 8.43** | 6 / 6 | 100.0 | **2.00** | **1.26, 1.67** | 5 / 6 | 83.3 | 1.00 | 0.66, 1.51 |
| Yes, fear of discrimination | 10 / 16 | 62.5 | **2.81** | **1.09, 7.23** | 13 / 16 | 81.2 | 1.62 | 0.97, 2.73 | 16 / 16 | 100.0 | 1.20 | 0.98, 1.47 |
| Yes, to avoid dysphoria or physical discomfort | 30 / 61 | 49.2 | 2.21 | 0.90, 5.45 | 43 / 61 | 70.5 | 1.41 | 0.86, 2.30 | 54 / 61 | 88.5 | 1.06 | 0.85, 1.33 |
| <u>Have received any shots of the HPV vaccine</u> |  |  |  |  |  |  |  |  |  |  |  |  |
| No | 18 / 43 | 41.9 | 0.81 | 0.53, 1.24 | 26 / 43 | 60.5 | 0.78 | 0.59, 1.03 | 36 / 43 | 83.7 | 0.90 | 0.78, 1.04 |
| Yes | 30 / 58 | 51.7 | 1.00 | (Ref) | 45 / 58 | 77.6 | 1.00 | (Ref) | 54 / 58 | 93.1 | 1.00 | (Ref) |
| <u>Using the anal swab was not difficult</u> |  |  |  |  |  |  |  |  |  |  |  |  |
| No | 4 / 30 | 13.3 | **0.21** | **0.08, 0.54** | - | - | - | - | 21 / 30 | 70.0 | **0.72** | **0.57, 0.91** |
| Yes | 44 / 71 | 62.0 | 1.00 | (Ref) | - | - | - | - | 69 / 71 | 87.2 | 1.00 | (Ref) |
| <u>Using the anal swab was not uncomfortable</u> |  |  |  |  |  |  |  |  |  |  |  |  |
| No | - | - | - | - | 27 / 53 | 50.9 | **0.55** | **0.42, 1.73** | 42 / 53 | 79.3 | **0.79** | **0.69, 0.91** |
| Yes | - | - | - | - | 44 / 48 | 91.7 | 1.00 | (Ref) | 48 / 48 | 100.0 | 1.00 | (Ref) |
| <u>Preferred method of screening for anal cancer</u> |  |  |  |  |  |  |  |  |  |  |  |  |
| HPV self-test at home | 41 / 67 | 61.2 | 1.00 | (Ref) | 57 / 67 | 85.1 | 1.00 | (Ref) | 65 / 67 | 97.0 | 1.00 | (Ref) |
| HPV test in clinic collected by medical professional | 4 / 13 | 30.8 | 0.50 | 0.22, 1.16 | 4 / 13 | 30.8 | **0.36** | **0.16, 0.82** | 10 / 13 | 76.9 | 0.79 | 0.59, 1.07 |
| Pap test in clinic | 2 / 8 | 25.0 | 0.41 | 0.12, 1.38 | 3 / 8 | 37.5 | 0.44 | 0.18, 1.08 | 6 / 8 | 75.0 | 0.77 | 0.52, 1.16 |
| No preference | 1 / 13 | 7.7 | 0.13 | 0.02, 0.83 | 7 / 13 | 53.8 | 0.63 | 0.38, 1.06 | 9 / 13 | 69.2 | 0.71 | 0.49, 1.03 |

Overall, most participants reported they would be willing to use the anal self-swab for future anal HPV testing (89.1%). Participants were more likely to be willing to use the anal swab if they were older (40–59 vs 21–29 years old: PR 1.12; 95% CI 1.03, 1.22), resided in a rural area compared to a suburban area (PR 1.19; 95% CI 1.06, 1.35), if they had not visited a medical professional in the past year (PR 1.13; 95% CI 1.05, 1.22), and they had transitioned but were not currently using hormones for masculinization. Both finding the anal swab uncomfortable (PR 0.72; 95% CI 0.57, 0.91) and finding the anal swab difficult (PR 0.79; 95% CI 0.69, 0.91) were associated with decreased willingness to use the swab in the future. There was an overall preference for at-home self-swab for anal HPV testing, with 66.3% of the population preferring at home HPV testing, compared to 12.9% preferring an HPV test performed in clinic and collected by a medical professional, 7.9% preferring an anal Pap test, and 12.9% having no preference.

### Self-reported qualitative reasons for cerivcovaginal and anal swab difficulty

Fourteen participants (14%) reported a difficult and/or distressing experience with using the cervicovaginal swab, but ten (71%) of whom reported they would still be willing to use the cervicovaginal self-swab for HPV testing in the future. Among the four participants who gave reasons for their negative experience using the cervicovaginal swab, three cited pain or discomfort when using the swab, and two cited emotional distress or dysphoria. Discomfort and emotional distress were known potential risks disclosed in the consent form, and participants were given contact information for TGD-supportive resources. Thirty participants (30%) reported a difficult or distressing experience with using the anal swab, but twenty-one (70%) reported they would still be willing to use the anal self-swab for HPV testing in the future. Seventeen participants described their negative experiences. Nine participants described difficulty with inserting the swab into their anus, in some cases because of the size or shape of their bodies and needing partner assistance. Three participants described the experience as awkward or unfamiliar but not necessarily painful. Two reported emotional distress but noted that they appreciated the ability to do the test in the comfort of their own homes.

## Discussion

Our analysis of the perceptions of the cervicovaginal and anal self-swab collection in a Midwestern TGD population indicated an overall broad acceptance of self-swab collection methods for HPV testing. For both the cervicovaginal and anal swabs, most participants indicated they would be willing to use these self-collection methods again in the future for HPV testing and cancer screening (96% for cervicovaginal swab, 89% for anal swab). We also found that despite difficulty or discomfort, participants reported an overall willingness to use the swabs in the future (71% and 70% of those reporting difficulty/discomfort, respectively). The preference for at-home self-swab collection for HPV testing also suggests that self-collection may be a more acceptable and accessible option for TGD individuals, who may experience discomfort or discrimination in healthcare settings. Most demographic characteristics were not significantly associated with participants’ willingness to use the cervicovaginal self-swab for future cervicovaginal HPV testing. However, older and rural participants were more willing to use the anal self-swab.

While participants overall reported a positive experience with cervicovaginal swab self-sampling, a small fraction did express some discomfort with using the swab or reported that the process generated feelings of dysphoria and emotional distress. Several participants reported that although the act of cervicovaginal self-sampling was not necessarily inherently unpleasant or painful, it was awkward and outside of their usual experiences. Increased familiarity and normalization of the procedure may improve overall acceptability and comfort. Participants reported mixed experiences with the anal self-swab in particular. Some individuals encountered difficulty with insertion, with a few noting challenges in maneuvering the swab due to their body size and shape or lack of flexibility. Others required assistance from their partner to properly use the swab. These experiences suggest that alternative instructions or guidance for diverse body types and positions may be beneficial for future self-sampling protocols. Additionally, anxiety emerged as a common theme among participants for both the cervicovaginal and anal swabs. While some individuals expressed a sense of unease or emotional distress during the self-sampling processes, they also noted a preference for experiencing these emotions in the privacy of their homes rather than in a clinical setting. This result highlights the potential benefit of self-sampling in mitigating the anxiety that some individuals may experience during in-person screening procedures.

The findings of our study align with previous research on HPV testing within the TGD population, which has highlighted the unique challenges and barriers faced by this population.^5,9^ Most participants in our study reported delaying medical care due to concerns related to their gender identity. This result emphasizes the importance of addressing TGD-specific barriers to healthcare by improving provider knowledge in TGD healthcare and creating more inclusive and comfortable environments.^22^ The acceptance and willingness to use self-collection methods aligns with previous research suggesting that self-sampling for HPV testing is a promising alternative for TGD individuals who may experience discomfort or anxiety during traditional clinician-collected sampling methods.^23^ There is also growing evidence of overall acceptability of self-sampling methods for HPV testing among the cisgender female population, suggesting the potential for HPV self-sampling to increase cervical cancer screening rates when compared to the standard of care pap test.^24^ Additionally, our findings are consistent with those reported by Maza et al., who also examined the acceptability of HPV self-sampling among transgender men in El Salvador.^25^ Both studies highlight the potential benefits of self-sampling in addressing barriers to accessing cervical cancer screening services for TGD populations.

Our research provides healthcare professionals with a better understanding on providing medical care to the TGD population. Currently, the FDA has not approved self-collection methods for use in routine HPV screening, requiring individuals AFAB to undergo specimen collection by a physician in a medical clinic. Approval and implementation of self-collection methods for HPV testing can increase accessibility to cervical cancer screening by removing the need for a pelvic exam, which may be a barrier for some TGD individuals who experience gender dysphoria or discomfort with their genitals.^26^ While the preference is for patients to receive in-person healthcare services with a medical professional, we acknowledge the unique barriers faced by TGD and the need for gender-affirming environments in healthcare.^27^ We also acknowledge a profusion of internet-based services offering a variety of HPV testing; these tests are not FDA approved and are of uncertain accuracy.^28^ It is imperative that a clinically valid, at-home tests are developed and approved.

Additionally, while the USPSTF has moved to using gender neutral language and specify anatomical features as opposed to sex, there are no transgender specific screening guidelines.^4,29^ Lack of transgender-specific guidelines and research has been shown as a limiting factor in providing appropriate healthcare, with many providers lacking knowledge on cervical cancer risk and developing a multitude of different strategies to manage care, which may not be evidence-based or appropriate for this population; potentially leading to inadequate or inappropriate care and further contributing to disparities in healthcare.^12,30^ The absence of clear guidelines may also affect the patient-provider relationship, with transgender individuals potentially feeling misunderstood or unsupported by healthcare providers who are not well-versed in their specific healthcare needs.^30^ By developing transgender-specific guidelines and integrating self-sampling options, when approved, into cervical cancer screening protocols, healthcare providers can cater to the diverse needs of transgender patients, promote health equity, and ultimately improve outcomes in this underserved population.^31^

A strength of this study is the attention to the under-researched area of HPV and cervical cancer screening among the TGD population, including nonbinary and other gender diverse individuals specifically. By investigating multi-modal screening methods and focusing on a Midwestern cohort rather than urban, coastal areas, the study provides valuable insights into the broader TGD community’s preferences and experiences with self-sampling techniques. Additionally, this study incorporated feedback from members of the TGD community who were consulted during the study design to ensure that the study materials (consent, questionnaire, sampling instructions) was comprehensive and appropriate for the population. We collected both quantitative and qualitative data, which allowed for a comprehensive understanding of participants’ preferences and potential barriers to screening, which can inform future interventions and guidelines. Our study was limited in that the population sample size was relatively small (although typical of studies in TGD populations), which may limit the generalizability of the findings to the broader TGD population. Additionally, most participants identified as White, and thus our work may not adequately reflect the experiences and concerns of TGD individuals from other racial and ethnic backgrounds.

Our study demonstrated the overall acceptability cervicovaginal and anal self-sampling methods for HPV testing and cervical cancer screening among TGD individuals AFAB, and it lays the groundwork for future research and interventions aimed at addressing disparities in healthcare access and outcomes. Most participants reported a positive experience with the self-sampling methods, as well as an overall willingness to use these methods in the future despite any perceived difficulty or discomfort; these results may be indicative of a potential avenue to increase screening uptake and reduce barriers to healthcare in this underserved population. This study also highlights the need for tailored interventions for TGD individuals that address diverse preferences and experiences, and the need to develop evidence-based, transgender-specific guidelines for HPV testing and cervical cancer screening^25,32^. With continued research into the development of evidence-based and patient-centered guidelines and interventions tailored to the broader TGD population, we can work towards increasing accessibility and utilization of preventative healthcare services by TGD individuals.

## Data Availability

All data produced in the present study are available upon reasonable request to the authors. A data use agreement will be required.

## Acknowledgments

We acknowledge our community consultants and our participants, without whom this work would not have been possible.

## References

1. Cancers caused by HPV. Centers for Disease Control and Prevention. https://www.cdc.gov/hpv/parents/cancer.html. Published February 28, 2022. Accessed March 23, 2023.

2. Hirth J. Disparities in HPV vaccination rates and HPV prevalence in the United States: A review of the literature. Human Vaccines & Immunotherapeutics. 2018;15(1). Doi:10.1080/21645515.2018.1512453

3. Cervical cancer: Screening. Recommendation: Cervical Cancer: Screening | United States Preventive Services Taskforce. https://www.uspreventiveservicestaskforce.org/uspstf/recommendation/cervical-cancer-screening. Published August 21, 2018. Accessed March 23, 2023.

4. Dhillon N, Oliffe JL, Kelly MT, Krist J. Bridging barriers to cervical cancer screening in transgender men: a scoping review. American journal of men’s health. 2020 May;14(3):1557988320925691.

5. Kattari SK, Gross EB, Harner V, et al. “Doing It On My Own Terms”: Transgender and nonbinary adults’ experiences with HPV self-swabbing home testing kits. Women’s Reproductive Health. June 2022. Doi:10.1080/23293691.2022.2094737

6. James SE, Herman JL, Rankin S, Keisling M, Mottet L, Anafi M. The Report of the 2015 U.S. Transgender Survey. National Center for Transgender equality. https://transequality.org/sites/default/files/docs/usts/USTS-Full-Report-Dec17.pdf. Published December 2016. Accessed March 24, 2023.

7. Peitzmeier SM, Reisner SL, Harigopal P, Potter J. Female-to-male patients have high prevalence of unsatisfactory paps compared to non-transgender females: Implications for cervical cancer screening. Journal of General Internal Medicine. 2014;29(5):778–784. Doi:10.1007/s11606-013-2753-1

8. Gatos KC. A literature review of cervical cancer screening in transgender men. Nursing for women’s health. 2018 Feb 1;22(1):52–62.

9. Li Y, Xu C. Human papillomavirus-related cancers. Advances in Experimental Medicine and Biology. 2017:23–34. Doi:10.1007/978-981-10-5765-6_3

10. Brown B, Poteat T, Marg L, Galea JT. Human papillomavirus-related cancer surveillance, prevention, and screening among transgender men and women: Neglected populations at high risk. LGBT Health. 2017;4(5):315–319. Doi:10.1089/lgbt.2016.0142

11. Agénor M, Peitzmeier SM, Bernstein IM, et al. Perceptions of cervical cancer risk and screening among transmasculine individuals: Patient and provider perspectives. *Culture*, Health & Sexuality. 2016;18(10):1192–1206. Doi:10.1080/13691058.2016.1177203

12. Potter J, Peitzmeier SM, Bernstein I, et al. Cervical cancer screening for patients on the female-to-male spectrum: A narrative review and guide for Clinicians. Journal of General Internal Medicine. 2015;30(12). Doi:10.1007/s11606-015-3462-8

13. McRee A-L, Gower AL, Reiter PL. Preventive healthcare services use among transgender young adults. International Journal of Transgenderism. 2018;19(4):417–423. doi:10.1080/15532739.2018.1470593

14. Centers for Disease Control and Prevention. National Health and Nutrition Examination Survey (NHANES): HVP Rinse. https://www.n.cdc.gov/nchs/data/nhanes/2009-2010/manuals/HPV.pdf. Published 2009. Accessed March 24, 2023.

15. McIntosh RD, Andrus EC, Walling HM, Sandler CB, Goudsmit CM, Moravek MB, Stroumsa D, Kattari SK, Brouwer AF. Prevalence and determinants of cervicovaginal, oral, and anal HPV infection in a population of transgender and gender diverse people assigned female at birth. medRxiv.

16. Evalyn® Brush. Rovers Medical Devices. https://www.roversmedicaldevices.com/cell-sampling-devices/evalyn-brush/. Published October 7, 2022. Accessed April 20, 2023.

17. Nyitray AG, Carvalho da Silva RJ, Baggio ML, et al. Six-month incidence, persistence, and factors associated with persistence of anal human papillomavirus in men: The HPV in men study. Journal of Infectious Diseases. 2011;204(11):1711–1722. doi:10.1093/infdis/jir637

18. Patel P, Bush T, Kojic EM, et al. Prevalence, incidence, and clearance of anal high-risk human papillomavirus infection among HIV-infected men in the sun study. The Journal of Infectious Diseases. 2017;217(6):953–963. doi:10.1093/infdis/jix607

19. Eisenberg MC, Campredon LP, Brouwer AF, et al. Dynamics and determinants of HPV infection: The Michigan HPV and oropharyngeal cancer (M-hoc) study. BMJ Open. 2018;8(10). doi:10.1136/bmjopen-2018-021618

20. Gottschlich A, Rivera-Andrade A, Grajeda E, Alvarez C, Mendoza Montano C, Meza R. Acceptability of human papillomavirus self-sampling for cervical cancer screening in an indigenous community in Guatemala. Journal of Global Oncology. 2017;3(5):444–454. doi:10.1200/jgo.2016.005629

21. Zhu X, MacLaughlin KL, Fan C, et al. Awareness of HPV testing and acceptability of self-sampling for cervical cancer screening among women in Minnesota. Journal of General Internal Medicine. 2021;37(6). doi:10.1007/s11606-021-06854-x

22. Seay J, Ranck A, Weiss R, Salgado C, Fein L, Kobetz E. Understanding transgender men’s experiences with and preferences for cervical cancer screening: A rapid assessment survey. LGBT Health. 2017;4(4):304–309. doi:10.1089/lgbt.2016.0143

23. Reisner SL, Vetters R, Leclerc M, et al. Mental health of transgender youth in care at an adolescent urban community health center: A matched retrospective cohort study. Journal of Adolescent Health. 2015;56(3):274–279. doi:10.1016/j.jadohealth.2014.10.264

24. Johnson AH, Hill I, Beach-Ferrara J, Rogers BA, Bradford A. Common barriers to healthcare for transgender people in the U.S. Southeast. International Journal of Transgender Health. 2019;21(1):70–78. doi:10.1080/15532739.2019.1700203

25. Yeh PT, Kennedy CE, de Vuyst H, Narasimhan M. Self-sampling for human papillomavirus (HPV) testing: A systematic review and meta-analysis. BMJ Global Health. May 2019. doi:10.1136/bmjgh-2018-001351

26. Maza M, Meléndez M, Herrera A, et al. Cervical cancer screening with human papillomavirus self-sampling among transgender men in El Salvador. LGBT Health. 2020;7(4):174–181. doi:10.1089/lgbt.2019.0202

27. Connolly D, Hughes X, Berner A. Barriers and facilitators to cervical cancer screening among transgender men and non-binary people with a cervix: A systematic narrative review. Preventive Medicine. 2020;135. doi:10.1016/j.ypmed.2020.106071

28. Gender-affirming care for young people: A guide for healthcare providers. U.S. Department of Health and Human Services. https://opa.hhs.gov/sites/default/files/2022-03/gender-affirming-care-young-people-march-2022.pdf. Published 2022. Accessed March 24, 2023.

29. Underferth D. Should you get a home HPV test? MD Anderson Cancer Center. https://www.mdanderson.org/cancerwise/should-you-get-a-home-hpv-test.h00-159463212.html. Published August 18, 2021. Accessed April 15, 2023.

30. Caughey AB, Krist AH, Wolff TA, et al. USPSTF approach to addressing sex and gender when making recommendations for Clinical Preventive Services. JAMA. 2021;326(19). doi:10.1001/jama.2021.15731

31. Snelgrove JW, Jasudavisius AM, Rowe BW, Head EM, Bauer GR. “completely out-at-sea” with “Two-gender medicine”: A qualitative analysis of physician-side barriers to providing healthcare for transgender patients. BMC Health Services Research. 2012;12(1). doi:10.1186/1472-6963-12-110

32. Poteat T, German D, Kerrigan D. Managing uncertainty: A grounded theory of stigma in transgender health care encounters. Social Science & Medicine. 2013;84:22–29. doi:10.1016/j.socscimed.2013.02.019

33. Reisner SL, Poteat T, Keatley JA, et al. Global health burden and needs of transgender populations: A Review. The Lancet. 2016;388(10042):412–436. doi:10.1016/s0140-6736(16)00684-x

